# Utility of sentinel surveillance as an early warning system for emerging pathogens in a low-resource setting: correlations of dengue, chikungunya, Zika, and COVID-19 trends between sentinel and passive surveillance systems in Puerto Rico, 2012–2023

**DOI:** 10.1101/2025.03.07.25323522

**Authors:** Alfonso C. Hernandez-Romieu, Zachary J. Madewell, Mark Delorey, Hannah R. Volkman, Liliana Sanchez-Gonzalez, Vanessa Rivera-Amill, Diego Sainz, Jorge Bertrán-Pasarell, Verónica M. Frasqueri-Quintana, Jomil Torres Aponte, Melissa Marzan-Rodríguez, Aidsa Rivera, Olga Lorenzi, Carla P. Espinet-Crespo, Yashira Maldonado, Roberta Lugo-Robles, Gilberto A. Santiago, Janice Perez-Padilla, Jorge L. Muñoz-Jordán, Gabriela Paz-Bailey, Laura E. Adams

## Abstract

The representativeness and timeliness of sentinel surveillance for endemic and emerging arboviral and respiratory diseases in low-resource settings are understudied. We compared laboratory-confirmed epidemic dengue, non-epidemic dengue, Zika, chikungunya, and COVID-19 (pre-Omicron and Omicron periods) cases reported in Puerto Rico’s Sentinel Enhanced Dengue Surveillance System (SEDSS) with island-wide trends reported by the Department of Health’s passive disease surveillance system (PADSS). We plotted trends over time to assess representativeness and used lagged cross-correlations to determine whether SEDSS reporting preceded PADSS. SEDSS trends were representative of island-wide trends for all pathogens. SEDSS preceded reporting in PADSS by up to three, eight, and two weeks for epidemic dengue, Zika, and pre-Omicron COVID-19, respectively. Increasing case trends for non-epidemic dengue and chikungunya occurred simultaneously in both systems. In Puerto Rico, sentinel surveillance was representative of island-wide trends and could provide early warning for dengue epidemics and emerging diseases, such as Zika, and COVID-19.

## Introduction

The COVID-19 pandemic and expansion of dengue and chikungunya in the Americas highlight the need to improve surveillance for emerging diseases.(1, 2) Arboviral and respiratory viral pathogens have unpredictable and rapid transmission dynamics, requiring surveillance systems that can accurately quantify cases, identify changes in trends in real time, and provide early warning of increasing transmission.(3–11) Many low-resource settings rely on passive surveillance that is limited by case underreporting, incomplete laboratory confirmation, inconsistent epidemiologic and clinical data, and reporting delays.(6, 12) Although more resource intensive, sentinel surveillance systems can be implemented in key locations and may be more sensitive in detecting emerging and endemic arboviral and respiratory pathogens.

Sentinel infectious disease surveillance systems are designed to detect emerging pathogens, characterize epidemic trends, and serve as early warning systems in public health. Unlike passive surveillance, sentinel surveillance systems leverage consistent and robust testing of a defined population to detect pathogens more quickly. These systems are not reliant on clinical suspicion to trigger diagnostic testing and are not subject to receipt of information from multiple reporting entities, both of which can limit the completeness and timeliness of passive surveillance test results. Sentinel surveillance systems have proven effective in detecting and characterizing outbreaks of dengue, malaria, HIV/AIDS, and other infectious causes of acute febrile and respiratory illnesses.(13–15). There are limited data, however, regarding the use of sentinel surveillance to obtain representative and timely estimates of emerging arboviral and respiratory pathogen transmission.(16–19) We compared Puerto Rico’s Sentinel Enhanced Dengue Surveillance System (SEDSS) and the Puerto Rico Department of Health (PRDH) passive disease surveillance system (PADSS) to better understand the timeliness and representativeness of sentinel surveillance vis-à-vis passive surveillance for confirmed dengue, chikungunya, Zika, and COVID-19 cases reported during 2012–2023. Our objectives were to 1) determine the representativeness of trends reported in SEDSS compared to island-wide trends reported in PADSS, and 2) examine whether SEDSS could act as an early warning system by identifying trends in cases before PADSS. Our analyses can inform whether a sentinel surveillance system like SEDSS can serve as a reliable and timely surveillance proxy for estimating pathogen transmission trends in settings where under-resourced public health systems may face compounding challenges in the disproportionate disease burden.

## Methods

### Study design

We compared cases of laboratory-confirmed epidemic dengue (defined as periods with high transmission rates, 2012–2014), non-epidemic dengue (defined as periods with lower or sporadic transmission rates, 2019–2023), Zika (2016–2017), chikungunya (2014–2015), and COVID-19 (during pre-Omicron and Omicron periods) between SEDSS and PADSS. We did not include 2015–2018 in the non-epidemic dengue period due to very low case counts (total n=237 for the 4 years) that were insufficient to provide meaningful insights into dengue virus transmission patterns.

### Sentinel Enhanced Dengue Surveillance System (SEDSS)

Patients with acute febrile and respiratory illnesses presenting at emergency departments (ED) and outpatient clinics in Puerto Rico are offered enrollment in SEDSS. (20–22) SEDSS operated primarily from the Ponce region during 2012–2017 and expanded to include the San Juan Metro Area in 2018.(20) SEDSS began in May 2012 at Saint Luke’s Episcopal Hospital, a 427 inpatient bed tertiary care hospital that receives ∼60,000 patients/year and is the regional pediatric referral hospital for the Ponce and Mayaguez (∼500,000 residents) Health Districts. The Outpatient Acute Care Clinic (2016-present) provides services to ∼20,000 patients/year. Saint Luke’s Guayama (2013-2015), now known as Guayama Menonita Hospital, is a 161-bed secondary care hospital site that receives ∼35,000 patients per year from southeastern Puerto Rico (∼50,000 residents). Auxilio Mutuo Hospital, which serves the San Juan Metro area, began participating in SEDSS in 2018 and is a 500 inpatient bed tertiary care hospital with ∼40,000 emergency room visits and ∼14,500 admissions annually.

Patients reporting fever at presentation or in the last 7 days were offered enrollment in SEDSS. Patients could participate once per 14-day period. During the Zika epidemic (June 2016–June 2018), patients with rash, arthritis, arthralgia, or conjunctivitis were eligible regardless of fever. Starting in April 2020, patients with cough or shortness of breath in the last 14 days (with or without fever) were also eligible. Participant serum specimens were tested for dengue virus (DENV) 1–4 (from 2012), chikungunya virus (CHIKV) (from May 2014), and Zika virus (ZIKV) (from November 2015) using reverse transcription-polymerase chain reaction (RT-PCR) and IgM antibody capture (MAC)-ELISA. Before March 2016, DENV was detected using the CDC DENV-1–4 RT-PCR assay.(23) From March 2016–April 2020, the CDC Trioplex RT-PCR assay was used to detect DENV, CHIKV, and ZIKV simultaneously,(24) with DENV serotype confirmed using the CDC DENV-1–4 RT-PCR assay. After April 2020, the CDC DENV-1–4 RT-PCR assay was again used as the primary test for DENV. Participants also provided a nasopharyngeal swab for molecular testing for SARS-CoV-2 (from March 2020), influenza A and B, respiratory syncytial virus, parainfluenza 1–4, rhinovirus, human metapneumovirus, and human adenovirus by RT-PCR (from 2012).(25)

SEDSS enrolls a relatively consistent number of participants per week, with annual averages ranging from 35 participants per week in 2012 to 121 participants per week in 2019. Across all years, the overall weekly average is approximately 80 participants (median: 74, IQR: 53–103). Weekly enrollment variability is primarily influenced by the number of eligible patients presenting to care during enrollment hours and staffing levels. The median participation rate ranges from 25–33% in tertiary hospitals and 45–52% in the urgent care clinic. Drop-out rates are <1%. General metrics on participant eligibility, recruitment, and testing for SEDSS sites during 2012–2022 are detailed elsewhere.(20)

### Puerto Rico Department of Health (PRDH) passive surveillance system (PADSS)

As mandated by territorial and federal regulations, all positive results from DENV, CHIKV, and ZIKV RT-PCR and IgM antibody tests, and SARS-CoV-2 RT-PCR and antigen tests conducted in Puerto Rico during the study period had to be reported to PADSS. Suspected infections from symptomatic cases were reported to PADSS based on clinical suspicion, followed by laboratory confirmation. PADSS offers public access to de-identified, individualized test and outcome data for COVID-19 through a web-based dashboard and data repository.

### Definitions

For both PADSS and SEDSS, epidemic dengue cases were identified by the positive detection of DENV by PCR or IgM tests during January 1, 2012 – December 31, 2014. Non-epidemic dengue cases included positive PCR results or IgM positive results for DENV during January 1, 2019 – December 31, 2023. We considered IgM results for ZIKV where available in addition to positive dengue IgM results, to account for notable cross-reactivity between dengue IgM and Zika IgM antibodies in serologic tests. Cases with positive DENV IgM results but without ZIKV IgM testing after Zika IgM testing ceased in April 2020 were also included in the analysis, acknowledging the unavailability of ZIKV IgM testing during that period.

CHIKV cases were defined as those with positive tests during May 28, 2014 July 31, 2015. ZIKV cases were those from January 21, 2016, through March 23, 2017. Chikungunya and Zika cases were defined by positive PCR or IgM test results. Zika IgM included negative IgM results for DENV. We restricted analyses of CHIKV and ZIKV to the time periods when the viruses were first detected by either surveillance system through dates covering 95% of the cases identified during the study period. The later stages of the outbreaks were excluded from our analysis to provide a more focused assessment of the surveillance systems’ effectiveness in detecting cases during the peak epidemic phases.

COVID-19 cases in SEDSS are defined as any SEDSS participant who tested positive for SARS-CoV-2 by RT-PCR during a single illness event during March 8, 2020–February 28, 2022. Cases prior to November 28, 2021 (i.e., the date the first case of the Omicron variant was reported in Puerto Rico) were categorized as pre-Omicron, while cases on or after this date were categorized as Omicron. We stratified into pre-Omicron and Omicron periods due to an increase in SARS-CoV-2 testing through PADSS at the emergence of the Omicron variant. SEDSS cases and tests for all pathogens, which are reported to PADSS, were excluded from case and test counts to prevent double counting of individual illness events.

### Statistical Analysis

We described frequencies of laboratory confirmed cases of epidemic (2012–2014) and non-epidemic (2019–2023) dengue, chikungunya (2014–2015), Zika (2016–2017), and COVID-19 (2020–2022) by surveillance system, health region, and type of laboratory test. We calculated median and interquartile ranges (IQR) from symptom onset to specimen collection and from specimen collection to testing by virus and surveillance system. Comparisons of frequencies and medians were performed using chi-square and Wilcoxon test statistics.

To visually determine the representativeness of SEDSS relative to PADSS we plotted weekly case counts of DENV, CHIKV, ZIKV, and COVID-19 with smoothed density plots using kernel density estimates. The smoothing bandwidths (or standard deviations of the smoothing kernel) were based on Silverman’s rule of thumb. The joint bandwidths for DENV (2012–2014), DENV (2019–2023), CHIKV, ZIKV, and COVID-19 were 34.8, 52.8, 20.3, 8.4, and 28.8 days, respectively. We used the symptom onset date rather than the specimen collection date to summarize weekly counts as it more accurately reflects when pathogen transmission is occurring.

To examine whether SEDSS could act as an early warning system by identifying trends in cases before PADSS, we lagged weekly data from SEDSS to PADSS by +/- 12 weeks. We evaluated the cross-correlation function (CCF) at each lag to determine at which lagged week SEDSS cases were most correlated with week 0 for PADSS (i.e., data available to public health authorities when monitoring case trends). A CCF approaching 1.0 indicates high agreement. For example, if the highest CCF was observed at week -2 in SEDSS relative to PADSS, this would indicate that case reports in SEDSS preceded PADSS by two weeks thus offering some anticipated warning. To make this determination less empirical, models were fit to SEDSS and PADSS weekly case numbers using the `tscounts` package and we computed Anscombe residuals for both models. We then drew a bootstrap sample from the residuals from each of the two sets, and we paired the residual time series for further analysis. We next transformed these residual series back to the original (observed) scale and computed the CCF and the differences in the CCF between lags. We repeated this 5,000 times using the 2.5 and 97.5 percentiles of these replicates for confidence bounds. This allowed us to infer with some confidence whether the value of the CCF at a given lag was statistically higher from the value of the CCF at another lag. This was done for each virus period (DENV 2012–2014, DENV 2019–2023, CHIKV 2014–2015, ZIKV 2016–2017, SARS-CoV-2 2020–2022). Because SARS-CoV-2 antigen tests were not performed in SEDSS, only RT-PCR tests were used to calculate cross-correlations between SEDSS and PADSS. All analyses were done using R software, version 4.4.1 (R Foundation for Statistical Computing, Vienna, Austria).

### Ethics statement

The Centers for Disease Control and Prevention (CDC) and Ponce Medical School Foundation (PMSF) Institutional Review Boards approved the study protocol.

## Results

### Cases of dengue, chikungunya, Zika, and COVID-19 in SEDSS and PADSS

Number of cases, including their geographic distribution, and median times from symptom onset to specimen collection and from specimen collection to testing for all pathogens are shown in Table 1. Large differences were observed in the median time from specimen collection to PCR testing between the two systems, with SEDSS providing significantly faster turn-around times for all studied pathogens, except for COVID-19, where times from specimen collection to testing were not available. The median time from specimen collection to RT-PCR testing by pathogen in SEDSS and PADSS, respectively, were 11- 15 days for epidemic dengue, 7 and 8 days for non-epidemic dengue, 68 and 166 days for chikungunya, and 4 and 14 days for Zika (P-values ≤0.01). Median times were also significantly lower is SEDSS compared to PADSS for IgM testing for epidemic dengue (9 vs 14 days) and Zika (12 and 17 days)(P-values ≤0.01). With the exception of COVID-19, the majority of cases reported in SEDSS were from the Ponce Region, while the majority of cases reported in PADSS had a wider distribution in the San Juan Metro Area.

**Table 1.** Confirmed cases of DENV in epidemic and non-epidemic periods, CHIKV, ZIKV, and COVID-19 in pre-Omicron and Omicron periods reported in SEDSS and PADSS, Puerto Rico, United States.

|  | DENV<br>epidemic period<br>(2012–2014) |  | DENV<br>non-epidemic period<br>(2019–2023) |  | CHIKV (2014–2015) |  | ZIKV (2016–2017) |  | COVID-19<br>pre-Omicron<br>(2020–2021) |  | COVID-19<br>Omicron<br>(2022) |  |
| --- | --- | --- | --- | --- | --- | --- | --- | --- | --- | --- | --- | --- |
|  | SEDSS | PADSS | SEDSS | PADSS | SEDSS | PADSS | SEDSS | PADSS | SEDSS | PADSS | SEDSS | PADSS |
|  | n (%) | n (%) | n (%) | n (%) | n (%) | n (%) | n (%) | n (%) | n (%) | n (%) | n (%) | n (%) |
| **Laboratory confirmed cases, total\*** | 1126 | 16164 | 285 | 3234 | 2303 | 4650 | 1916 | 34995 | 544 | 188728 | 238 | 284601 |
| PCR | 738 (65.5) | 11120 (68.8) | 280 (98.2) | 3083 (95.3) | **1810 (78.6)** | **4417 (95.0)** | **1448 (75.6)** | **33764 (96.5)** | **544 (100)** | **155223 (82.2)** | **238 (100)** | **173433 (60.9)** |
| IgM<sup>†</sup> | **854 (75.8)** | **7511 (46.5)** | **89 (31.2)** | **1931 (59.7)** | **889 (38.6)** | **358 (7.7)** | **1255 (65.5)** | **3509 (10.0)** | NA | NA | NA | NA |
| SARS-CoV-2 antigen | NA | NA | NA | NA | NA | NA | NA | NA | 0 (0) | 33505 (17.8) | 0 (0) | 111168 (39.1) |
| Time from symptom onset to specimen collection<sup>‡</sup> | **3 (1, 4)** | **4 (3, 5)** | **3 (2, 4)** | **4 (2, 5)** | 1 (1, 2) | 1 (1, 2) | 2 (1, 3) | 2 (1, 4) | 2 (1, 3) | NA | 2 (1, 3) | NA |
| Time from specimen collection to PCR testing<sup>‡</sup> | **11 (8, 16)** | **15 (10, 26)** | **7 (5, 10)** | **8 (5, 16)** | **68 (29, 201)** | **166 (73, 261)** | **4 (3, 6)** | **14 (8, 22)** | 6 (3, 8) | NA | 5 (3, 7) | NA |
| Time from specimen collection to IgM testing<sup>‡</sup> | **9 (7, 13)** | **14 (9, 21)** | 12 (7, 19) | 13 (8, 25) | 76 (57, 104) | 72 (43, 103) | **12 (7, 23)** | **17 (8, 35)** | NA | NA | NA | NA |
| **Health Region§** |  |  |  |  |  |  |  |  |  |  |  |  |
| Aguadilla | 2 (0.2) | 1821 (11.3) | 0 (0) | 174 (5.4) | 2 (0.1) | 140 (3.0) | 0 (0) | 1816 (5.2) | 2 (0.4) | NA | 2 (0.8) | NA |
| Arecibo | 0 (0) | 1854 (11.5) | 2 (0.7) | 287 (8.9) | 4 (0.2) | 469 (10.1) | 2 (0.1) | 4050 (11.6) | 10 (1.8) | NA | 0 (0) | NA |
| Bayamon | 0 (0) | 2376 (14.7) | 20 (7.0) | 811 (25.1) | 5 (0.2) | 238 (5.1) | 5 (0.3) | 8067 (23.1) | 61 (11.2) | NA | 23 (9.7) | NA |
| Caguas | 15 (1.3) | 1133 (7.0) | 7 (2.5) | 263 (8.1) | 8 (0.3) | 1115 (24.0) | 6 (0.3) | 2950 (8.4) | 24 (4.4) | NA | 5 (2.1) | NA |
| Fajardo | 3 (0.3) | 286 (1.8) | 7 (2.5) | 54 (1.7) | 14 (0.6) | 738 (15.9) | 1 (0.1) | 380 (1.1) | 20 (3.7) | NA | 2 (0.8) | NA |
| Mayaguez | 6 (0.5) | 2745 (17.0) | 0 (0) | 307 (9.5) | 0 (0) | 161 (3.5) | 2 (0.1) | 3450 (9.9) | 0 (0) | NA | 3 (1.3) | NA |
| San Juan | 36 (3.2) | 3849 (23.8) | 196 (68.8) | 1216 (37.6) | 300 (13.0) | 1739 (37.4) | 14 (0.7) | 6683 (19.1) | 267 (49.1) | NA | 58 (24.4) | NA |
| Ponce | 1064 (94.5) | 2010 (12.4) | 52 (18.2) | 110 (3.4) | 1955 (84.9) | 32 (0.7) | 1885 (98.4) | 7464 (21.3) | 157 (28.9) | NA | 144 (60.5) | NA |
| Unknown | 0 (0) | 90 (0.6) | 1 (0.4) | 12 (0.4) | 15 (0.7) | 18 (0.4) | 1 (0.1) | 135 (0.4) | 3 (0.1) | NA | 1 (0.4) | NA |

DENV = Dengue Virus; CHIKV = Chikungunya Virus; ZIKV = Zika Virus; SARS-CoV-2 = Severe acute respiratory syndrome coronavirus 2; SEDSS = Sentinel Enhanced Dengue Surveillance System; PADSS Puerto Rico Dept of Health Passive Disease Surveillance System; PCR = Polymerase chain reaction; IgM = Immunoglobulin M; NA = not available (we were unable to access this data); IQR = Interquartile range
DENV epidemic period included positive tests from January 1, 2012, to December 31, 2014; DENV non-epidemic period: January 1, 2019, to December 31, 2023; CHIKV: May 28, 2014, to July 31, 2015; ZIKV: January 21, 2016, to March 23, 2017; COVID-19 : pre-Omicron March 8, 2020, to November 27, 2021, Omicron November 28, 2021 to February 28, 2022.
Bolded values indicate comparisons with P < 0.01.
\*Persons who underwent testing with both PCR and IgM were counted once, and the earliest specimen collection date was used.
†IgM testing was conducted with IgM antibody capture (MAC)-ELISA. During the epidemic period any IgM with a positive result was considered positive; during the non-epidemic period IgM with a positive result for DENV and a negative result for ZIKV were considered indicative of DENV infection.
‡Median (Interquartile range)
§ The metro region includes San Juan, Bayamon, and Caguas

### Representativeness of dengue, chikungunya, Zika, and COVID-19 trends

Crude (Figure 1) and smoothed density plots (Figure 2) of weekly case counts of dengue, chikungunya, Zika, and COVID-19 in SEDSS and PADSS followed similar distributions. Smoothed density plots show that Zika cases peaked earlier in SEDSS compared to PADSS.

**Figure 1.**
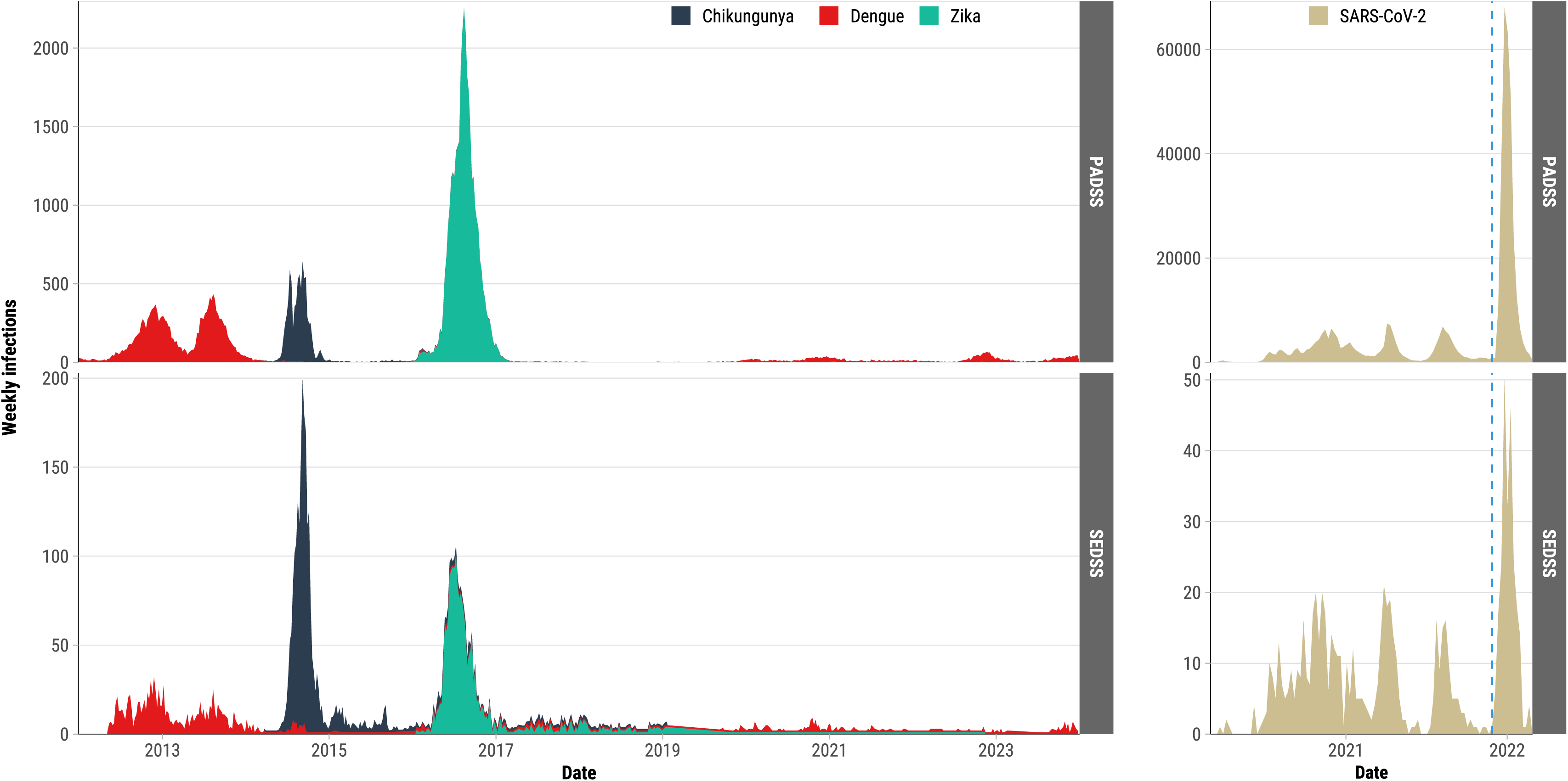
Weekly arbovirus and COVID-19 cases by symptom onset date and surveillance system type, Puerto Rico, 2012–2023. This figure compares cases reported by the Sentinel Enhanced Dengue Surveillance System (SEDSS) and the Puerto Rico Department of Health (PRDH) Passive Disease Surveillance System (PADSS). Arbovirus trends (left panels) include dengue, chikungunya, and Zika, while COVID-19 trends (right panels) are divided into pre-Omicron (before November 28, 2021) and Omicron (November 28, 2021, and after) periods. The dashed blue line indicates the beginning of the Omicron period.

**Figure 2.**
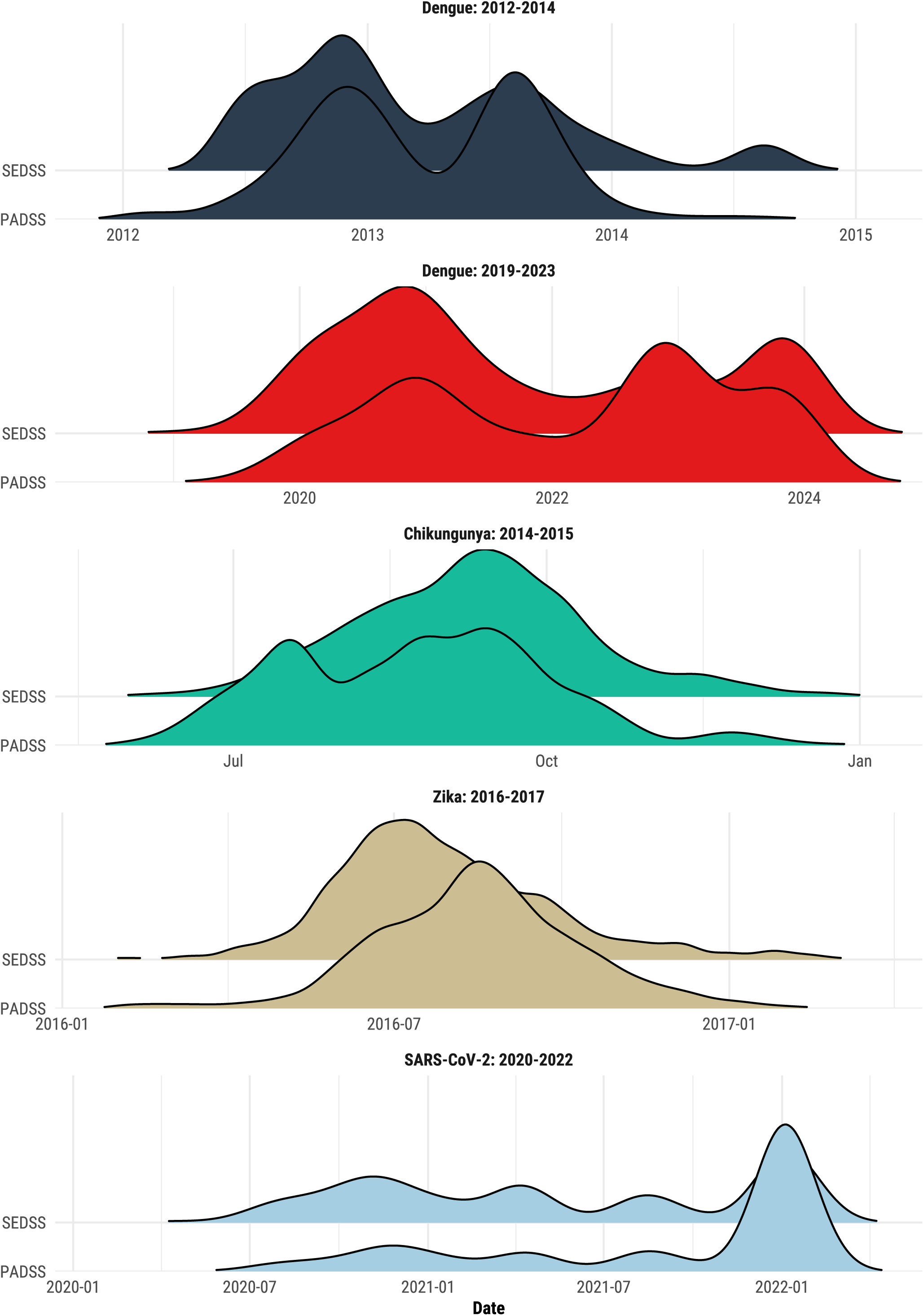
Smoothed kernel density plots of weekly arbovirus and COVID-19 cases by symptom onset date and surveillance system type, Puerto Rico, 2012–2023. This figure shows the distribution of cases reported by the Sentinel Enhanced Dengue Surveillance System (SEDSS) and the Puerto Rico Department of Health Passive Disease Surveillance System (PADSS) for dengue (2012–2014, 2019–2023), chikungunya (2014–2015), Zika (2016–2017), and COVID-19 (2020–2022). Each panel represents a specific pathogen and time period, whereas the y-axis separates the two surveillance systems. Kernel density estimates are smoothed to highlight temporal trends in case distributions, with joint bandwidths calculated for each pathogen. The time periods were defined using the following dates. DENV epidemic period: January 1, 2012–December 31, 2014; DENV non-epidemic period: January 1, 2019–December 31, 2023; CHIKV: May 28, 2014–July 31, 2015; ZIKV: January 21, 2016–March 23, 2017; COVID-19: pre-Omicron March 8, 2020–November 27, 2021; Omicron: November 28, 2021–February 28, 2022.

### Correlation of lagged dengue, chikungunya, Zika, and COVID-19 SEDSS cases to PADSS

During epidemic dengue the highest cross-correlation (0.77) was observed at a lag of 0 and -1 weeks (Table 2). The value of the CCF at lags 0 to -3 weeks were not statistically different (with 95% confidence) from that at its peak of 0.77. Thus, the data are consistent with a CCF whose peak actually occurs at a lag of somewhere from -3 to 0 weeks, inclusive, suggesting that SEDSS reporting precedes PADSS reporting by anywhere between 3 and 0 weeks. (Figure 3, panel A). The CCF for non-epidemic dengue was generally low, indicating low temporal concordance between the two reporting systems (Figure 4, panel B). Among chikungunya cases, the highest value of the CCF (0.90) was observed at week +2 and was not significantly different from lags at 0 and +1 weeks (Figure 4, panel C). The CCF for Zika was higher compared to other pathogens, reaching its highest value of 0.91 at -4 weeks. Lagged weekly Zika counts between 0 and -8 weeks were not significantly different, indicating that reported cases in SEDSS can precede reporting in PADSS by up to eight weeks. Among COVID-19 cases in the pre-Omicron period the highest CCF (0.82) occurred at lags of -1 and 0 weeks (Figure 4, panel E). CCFs at lags 0, -1, and -2 were not statistically different indicating that SEDSS cases may precede cases reported to PADSS by one or two weeks. In contrast, in the Omicron period, the highest CCF was observed at week 0 (0.93) and cross-correlations at lags –1, 0, and 1 were not statistically different from one another suggesting SEDSS cases were concordant but did not precede PADSS cases during the Omicron period.

**Figure 3.**
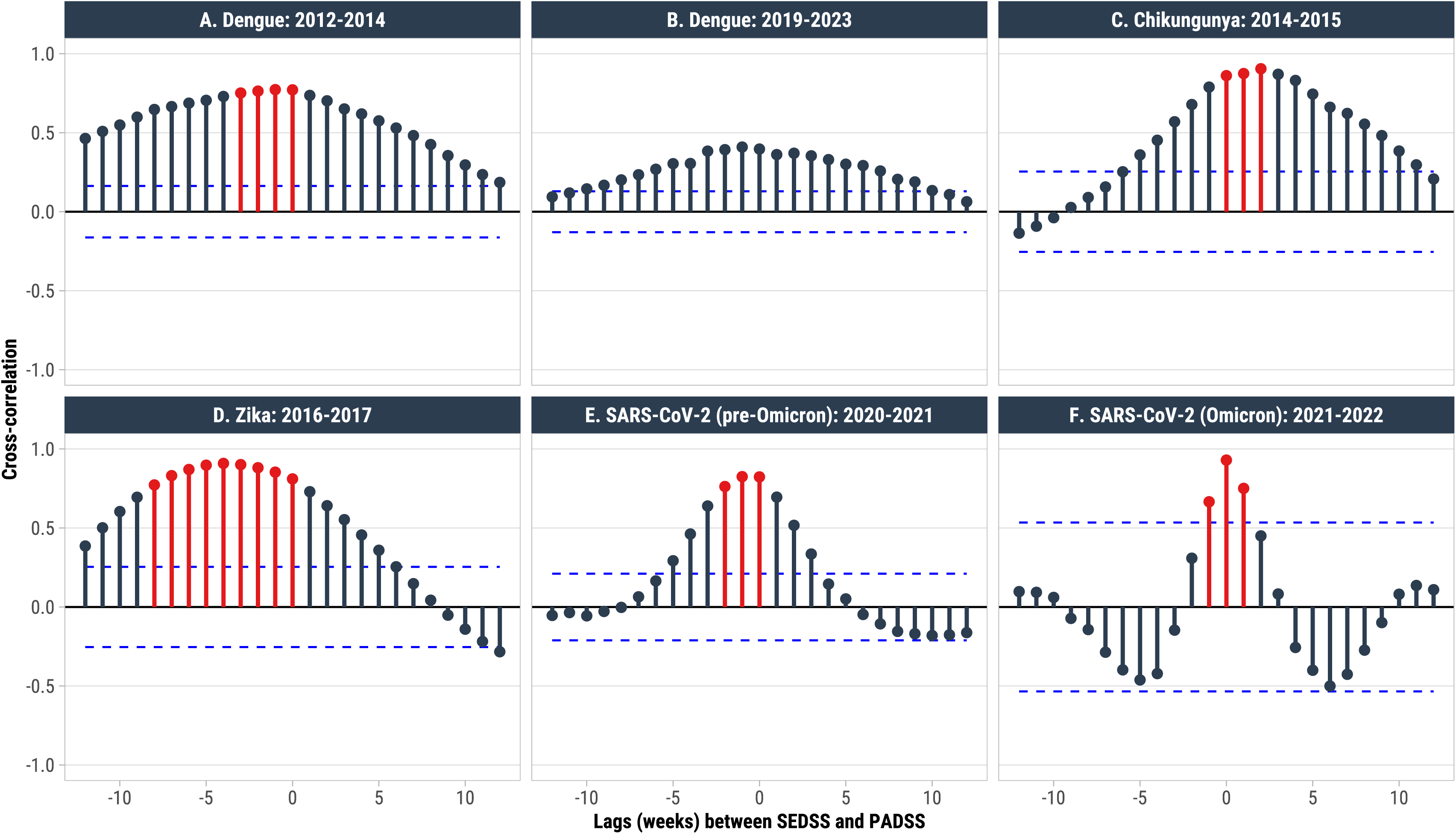
Cross-correlation coefficients (CCFs) comparing arboviral (panels A–D) and COVID-19 cases (panels E and F) between cases reported by symptoms onset date in SEDSS lagged +/- 12 weeks from PADSS. The blue dotted lines denote the significance threshold for individual CCFs. CCFs identified in red are not statistically different from each other, indicating that cases reported during those lags are similar. For example, in Panel A, epidemic dengue cases reported in SEDSS up to a lag of -3 weeks are highly correlated with PADSS at week 0 (i.e., when public health authorities should be receiving information on cases). This indicates that reporting in SEDSS precedes PADSS by up to three weeks. The time periods were defined using the following dates. DENV epidemic period: January 1, 2012–December 31, 2014; DENV non-epidemic period: January 1, 2019–December 31, 2023; CHIKV: May 28, 2014–July 31, 2015; ZIKV: January 21, 2016–March 23, 2017; COVID-19: pre-Omicron March 8, 2020–November 27, 2021; Omicron: November 28, 2021–February 28, 2022.

## Discussion

A sentinel surveillance system in Puerto Rico provided representative island-wide estimates and could serve as an early warning system for emerging and epidemic pathogens. Visual assessments between the sentinel and passive surveillance systems revealed similar trends for epidemic and non-epidemic dengue, chikungunya, Zika, and COVID-19. In addition, we observed that cases at sentinel sites could precede those in island-wide passive reporting by up to eight, three, and two weeks for Zika, epidemic dengue and pre-Omicron COVID-19, respectively. Reporting in SEDSS did not significantly precede PADSS for non-epidemic dengue cases; however, both systems tended to report cases simultaneously. Cases of chikungunya reported to PADSS may have preceded those in SEDSS by up to two weeks but were likely reporting cases at similar times as evidenced by the highest CCF at a week-zero lag. Our findings highlight the utility of sentinel surveillance in resource-limited settings, providing representative and timely estimates of transmission for emerging and epidemic pathogens.

Having a robust sentinel surveillance system in resource-limited settings has many advantages. First, SEDSS was able to rapidly implement testing for novel pathogens to detect ongoing circulation of Zika, chikungunya, and COVID-19. Despite significant delays during the chikungunya outbreak, times from specimen collection to testing were 50% shorter in SEDSS. Faster test implementation and turn-around times of specimens collected from a sentinel site during an outbreak of an emerging disease can enhance preparedness and monitoring of disease trends beyond the capabilities of a passive surveillance system. Second, standardized arboviral laboratory testing allowed differentiation of arboviral infections amidst overlapping signs and symptoms and co-circulation with respiratory viruses. Through SEDSS, we confirmed minimal circulation of dengue during the chikungunya and Zika epidemics. Third, during the COVID-19 pandemic, 59% of all PCR testing for dengue was performed in SEDSS, with <10% of participants testing positive, indicating low levels of dengue transmission that may have been overlooked if relying solely on the 30–60% test-positivity observed in the passive surveillance system. Finally, SEDSS allows for more precise denominators to measure hospitalization and complications of emerging viruses, such as chikungunya.

These findings also highlight the limitations of the sentinel surveillance approach. The first is that laboratory capacity can be rapidly overwhelmed in large epidemics. During the first year of the chikungunya epidemic, laboratory-confirmation of chikungunya was limited to hospitalized cases captured by PADSS and SEDSS. This limited the utility of the sentinel surveillance approach by biasing testing to more severe cases and decreasing the detection of more common, mild cases in a timely manner. A second limitation is related to the geographical variability of where cases occur. During the chikungunya epidemic, SEDSS was limited to the Ponce region. Although Ponce had a high incidence of chikungunya cases, the highest incidence was seen in the Metro San Juan Area and the northeastern region of the island. Thus, having limited capture of Puerto Rico’s population likely resulted in no advantage in case detection earlier than PADSS. In contrast, a SEDSS recruitment site was present in the San Juan Metro area during the COVID-19 pandemic which likely contributed to early warning for SEDSS relative to PADSS. Similarly, SEDSS failed to capture a large number of cases of non-epidemic dengue in 2023 from the western part of the island, which may have resulted in the low CCFs observed between SEDSS and PADSS. Enhancements to the geographical representativeness of sentinel sites could improve the early warning and monitoring of emerging infections.

Implementing sentinel surveillance for emerging pathogens requires careful consideration of multiple factors. First, costs can be high compared to passive surveillance in single facilities. Yearly direct and indirect costs to maintain multiple SEDSS sites, including laboratory supplies, staff, and study spaces, among others, amount to approximately $700,000 US dollars. However, costs may be significantly lower in other settings, and the preparedness and research capabilities of sentinel surveillance sites can be leveraged to offset costs to the funding entity through partnerships with academic institutions and public health agencies. Second, human and infrastructure needs are high. SEDSS requires dedicated recruiters, data analysts, and study site coordinators, as well as access to a reference laboratory that can promptly run tests and report back to study sites. Third, size matters. Given Puerto Rico’s size, 1-2 sentinel sites may be sufficient, whereas appropriate coverage for larger geographical areas would likely require more, thereby increasing costs. Finally, sentinel sites have lower limits of sensitivity and upper limits of recruitment, which may affect the generalizability of findings in large epidemics. During the Omicron period, SEDSS recruitment did not exceed 53 cases per week, which was not generalizable to the increased transmission observed during Omicron. Nevertheless, the benefits of early detection of transmission through sentinel surveillance, as observed in SEDSS, could significantly reduce morbidity, mortality, and economic costs that may offset initial investments. The findings from SEDSS were also shared in weekly reports to the Puerto Rico Department of Health and used during outbreaks to provide situational awareness and guide response planning decisions.

Our study is subject to at least four limitations. First, sentinel sites changed over time and they were not dispersed evenly throughout Puerto Rico, which may have affected the representativeness and timeliness of our findings and the variability by pathogen. However, the addition of a sentinel site in the San Juan Metro Area in 2018 likely increased the ability of SEDSS to detect emerging pathogens due to it’s high population density and being the port of entry into the island. Second, laboratory testing during the chikungunya outbreak of 2014–2015 was extremely delayed island-wide because of test availability. The 7-week anticipated chikungunya reporting in SEDSS compared to PADSS should be interpreted with caution. Third, SEDSS estimates are dependent on healthcare-seeking behavior. Participation in SEDSS is voluntary, and only those presenting to care can enroll. Fourth, relying primarily on hospital-based sentinel surveillance may have skewed our results, leading to under-capture of mild disease, although the comprehensive testing of all participants with fever could increase detection of dengue among patients with mild symptoms compared to surveillance systems where only hospitalized or severe cases are tested. Ensuring participation in outpatient clinics has been shown to provide better capture of mild-moderate disease during the COVID-19 pandemic and provide early warning relative to hospitalizations. Despite its limitations, our study’s strength stems from robust longitudinal data spanning multiple emerging pathogen epidemics and examination of different transmission periods for endemic dengue.

In summary, a sentinel site in Puerto Rico provided representative estimates of pathogen transmission and could serve as an early warning system for epidemics and emerging pathogens such as dengue, Zika, and SARS-CoV-2. Implementing sentinel surveillance in a low-resource setting should weigh the benefits of early detection of emerging pathogens against the higher cost of surveillance. Additionally, the ability to implement rapid public health action should be considered to determine whether this type of surveillance suits the context and needs.

## Acknowledgements

### Disclaimer

The findings and conclusions in this report are those of the authors and do not necessarily represent the official position of the US Centers for Disease Control and Prevention.

## Data Availability

Data are available from the CDC management team (contact) for researchers who meet the criteria for access to confidential data.

## Notes

### Competing Interest Statement

The authors have declared no competing interest.

### Funding Statement

This research was funded by Centers for Disease Control and Prevention, grant numbers U01CK000473 and U01CK000580 (VRA).

### Author Declarations

The Institutional Review Boards at the Centers for Disease Control and Prevention (CDC), Auxilio Mutuo, and Ponce Medical School Foundation approved the SEDSS study protocols 6214, and 120,308-VR/2311173707, respectively.

